# Effect of booster vaccination against Delta and Omicron variants in Iceland

**DOI:** 10.1101/2022.02.26.22271509

**Authors:** Gudmundur L Norddahl, Pall Melsted, Kristbjorg Gunnarsdottir, Gisli H Halldorsson, Thorunn A Olafsdottir, Arnaldur Gylfason, Mar Kristjansson, Olafur T Magnusson, Patrick Sulem, Daniel F Gudbjartsson, Unnur Thorsteinsdottir, Ingileif Jonsdottir, Kari Stefansson

## Abstract

By the end of July 2021, the majority of the Icelandic population had received vaccination against COVID-19. In mid-July a wave of SARS-CoV-2 infections, dominated by the delta variant, spread through the population, followed by an omicron wave in December. A booster vaccination campaign was initiated to curb the spread of the virus. We estimated the risk of infection for different vaccine combinations using vaccination data from 276,028 persons and 963,557 qPCR tests for 277,687 persons. We measured anti-Spike-RBD antibody levels and ACE2-Spike binding inhibitory activity in 371 persons who received one of four recommended vaccination schedules with or without an mRNA vaccine booster. Overall, we found different antibody levels and inhibitory activity between recommended vaccination schedules, which was reflected in the observed risk of SARS-CoV-2 infections. We observed an increased protection following mRNA boosters, against both omicron and delta variant infections, although BNT162b2 boosters provided greater protection against omicron than mRNA-1273 boosters.

## Introduction

Vaccines directed against SARS-CoV-2 have proven effective to reduce the risk of severe disease and transmission of the virus^1–5^. The four vaccines used in Iceland were the BNT162b2 messenger RNA (mRNA) vaccine (Pfizer–BioNTech), mRNA-1273 vaccine (Moderna) and adenoviral vector vaccines ChAdOx1 (AstraZeneca) and AD26.COV2.S (Janssen).

Front line workers and residents of nursing homes were the first to be vaccinated, followed by a nationwide effort to vaccinate the entire adult population. Emerging data on side effects lead to a targeted use of different vaccines by age and sex with ChAdOx1 nCoV-19 being primarily given to persons over 45 of age.^6^

At the end of July 2021, with the emergence of the B.1.617.2 (delta) variant of concern (VOC) and a following rise in transmission, Israeli authorities approved the administration of a third dose (booster) of the BNT162b2 mRNA vaccine. This led to a 90% lower mortality from infections among booster dose recipients than among those who received two dose vaccination.^7–9^

54,438 Icelanders (22% of those eligible for vaccine at the time) received the recommended one dose of Ad26.COV2.S as their primary vaccination. Due to a high incidence of SARS-CoV-2 infections among those who received one dose of Ad26.COV2.S vaccine, the Chief epidemiologist in Iceland issued a recommendation of a booster dose of mRNA vaccine to all Ad26.COV2.S vaccine recipients in early August 2021, which has been shown to be effective in raising antibody levels.^10,11^ Following this, booster vaccinations were recommended to the general public regardless of the vaccine they had initially received.^12^

The first case of the B.1.1.529 (omicron) VOC in Iceland was confirmed on December 1, 2021, and spread rapidly, reaching 90% of daily confirmed cases by January 6, 2022.^13^ Although vaccine effectiveness of BNT162b2 is reported to be lower against the omicron variant, booster vaccination has been shown to substantially elevate the neutralizing activity against the variant.^14–18^

Here we report on the effect of vaccination on serum anti-Spike-RBD antibody levels and their inhibitory activity in well-defined groups of vaccine recipients matched for age and sex, with detailed information on vaccination status and COVID-19 diagnosis, following both recommended vaccination schedules for each of the four vaccines and additional booster vaccinations. Finally, we use data on all vaccinations (n=276,028 persons), all PCR tests (n=963,557 tests for n=277,687 persons) and persons diagnosed with COVID-19 (n=31,919 persons) in Iceland to estimate the effect of these vaccination schedules on the risk of being diagnosed with COVID-19.

## Results

First, participants were recruited in July 2021 for measurements of antibody and inhibition levels. Initially, four groups were drawn from persons vaccinated with their last dose of the primary vaccine schedule in May 2021. The groups were defined by the vaccine received, two doses of BNT162b2, ChAdOx1, or mRNA-1273, and those who received one dose of Ad26.COV2.S. An age and gender matched list was generated at random and participants invited for the study. This resulted in 196 participants (Table 1, Figure 1), who gave a single sample two months after receiving their last dose.

**Table 1.** Overall demographics broken down by vaccines. Time from vaccination is measured from final vaccination date to the date of sample collection.

| Vaccine | Comparison of recommended vaccine schedules |  |  |  |  | Comparison of mRNA booster vaccination |  |  |  |  |
| --- | --- | --- | --- | --- | --- | --- | --- | --- | --- | --- |
|  | Overall | Ad26.COV2.S | ChAdOx1-ChAdOx1 | BNT162b2-BNT162b2 | mRNA-1273-mRNA-1273 | Overall | Ad26.COV2.S pre-booster | BNT162b2-BNT162b2 | Ad26.COV2.S - BNT162b2 | Ad26.COV2.S-mRNA-1273 |
| Number (persons) | 196 | 31 | 62 | 52 | 51 | 158 | 99 | 59 | 33 | 66 |
| Female (%) | 145 (73%) | 26 (84%) | 42 (68%) | 35 (67%) | 42 (82%) | 97 (61%) | 64 (65%) | 33 (56%) | 20 (61%) | 44 (67%) |
| Age (years) | 51.7 | 53 | 50.2 | 52 | 52.5 | 37.2 | 37.2 | 37.1 | 36.8 | 37.3 |
| Time from vaccination (days) | 49.5 | 50.5 | 50.3 | 46.0 | 51.6 | 59.6 | 62.6 | 52.8 | 60.1 | 62.7 |
| **Vaccine schedule** |  |  |  |  |  |  |  |  |  |  |
| *No of doses* |  | 1 | 2 | 2 | 2 | - | - | - | - | - |
| *Time interval* |  | - | 3 months | 3 weeks | 4 weeks | - | - | - | - | - |
| **Measurements** |  |  |  |  |  |  |  |  |  |  |
| Antibody U/ml (SD) |  | 113 (108) | 969 (2132) | 850 (885) | 3724 (3112) |  | 167 (140) | 1887 (4796) | 3356 (2063) | 5432 (2434) |
| Inhibition U/ml (SD) |  | 2.79 (4.04) | 4.02 (7.97) | 12.1 (14.3) | 33.4 (25.8) |  | 3.87 (2.06) | 22.6 (22.3) | 18.8 (11.5) | 33.7 (20.5) |

**Figure 1.**
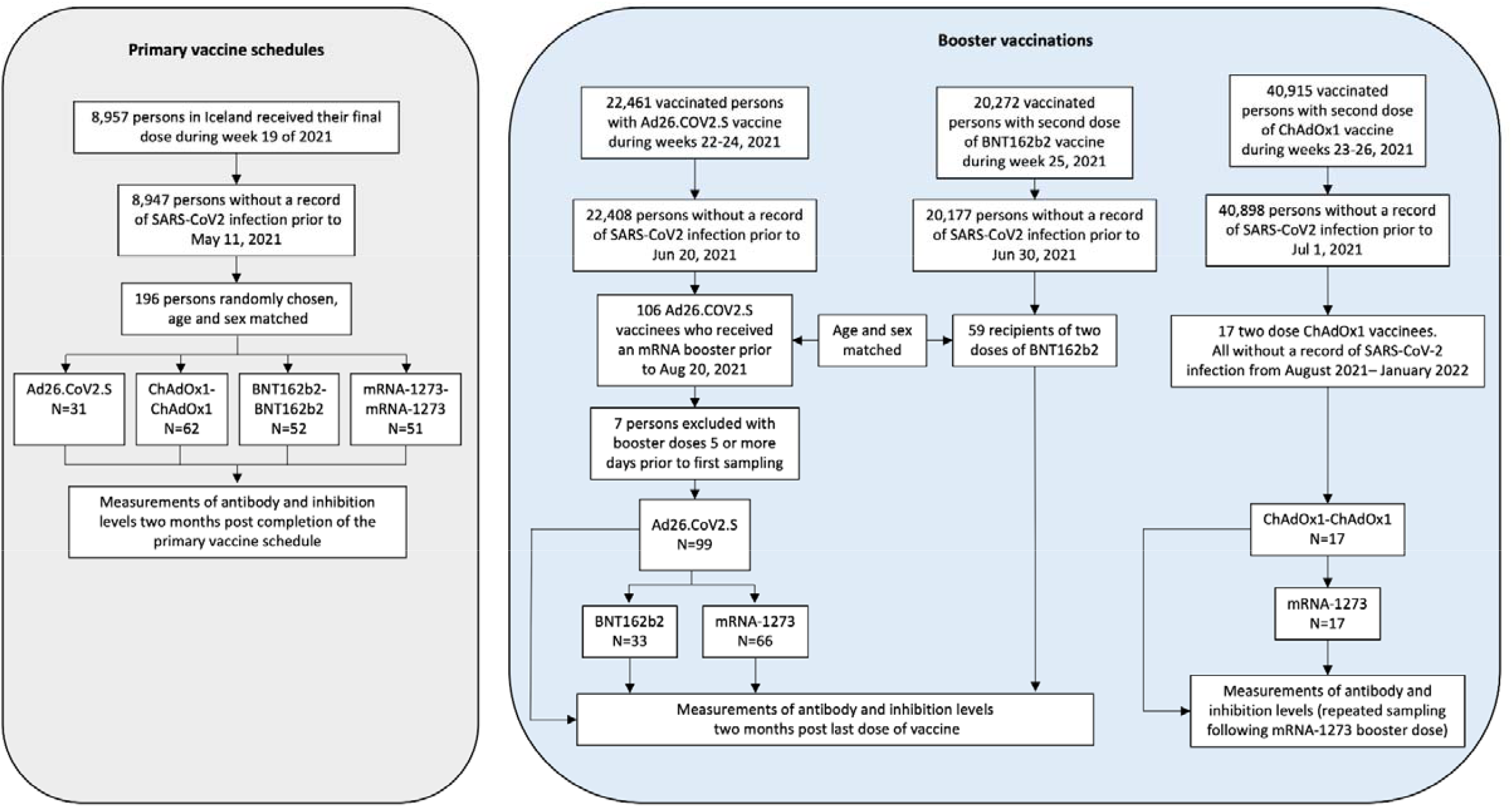
Schematic overview of the recruited groups used in the study.

In August 2021, a decision was made to offer persons who received a single dose of Ad26.COV2.S a booster dose of an mRNA vaccine. To study the effect on antibody and inhibition levels we recruited 106 persons who received a single dose of Ad26.COV2.S two months prior. Participants gave a sample prior to receiving a booster dose and a follow-up sample two months after. As this group differed in age composition compared to the initial study, we simultaneously recruited 59 persons, age and gender matched to the Ad26.COV2.S booster group, who received their second dose of BNT162b2 two months prior (Table 1, Figure 1).

To study waning of antibody and inhibition levels following a booster dose we recruited 17 persons in the beginning of August 2021, who received two doses of ChAdOx1 and a booster dose of mRNA-1273 (Table S1, Figure 1). The first sample was taken prior to receiving the booster dose and participants were followed up with frequent sampling over a period of 154 days.

Finally, to study the protection of vaccines against SARS-CoV-2 infections we conducted a retrospective total population cohort study in Iceland. We obtained data from the Vaccination Register (The Directorate of Health) and included all persons who had received at least one dose of a vaccine before January 8 2022 (n=278,026). For each person we obtained information about all vaccinations, the date of each dose administered and vaccine used, as well as age and gender (Table S2). We included only individuals aged 18-80 year old (Figure 2). We focused on two time periods, July 1 – November 22, 2021 and December 1 – January 8, 2022 that were dominated by delta and omicron infections respectively (Figure S1). We obtained data from the Register of Communicable Diseases (The Directorate of Health) on all PCR tests conducted in Iceland from January 30, 2020 to January 8, 2022 (n=963,557 tests for n=277,687 persons) to exclude all persons with a prior infection.

**Figure 2.**
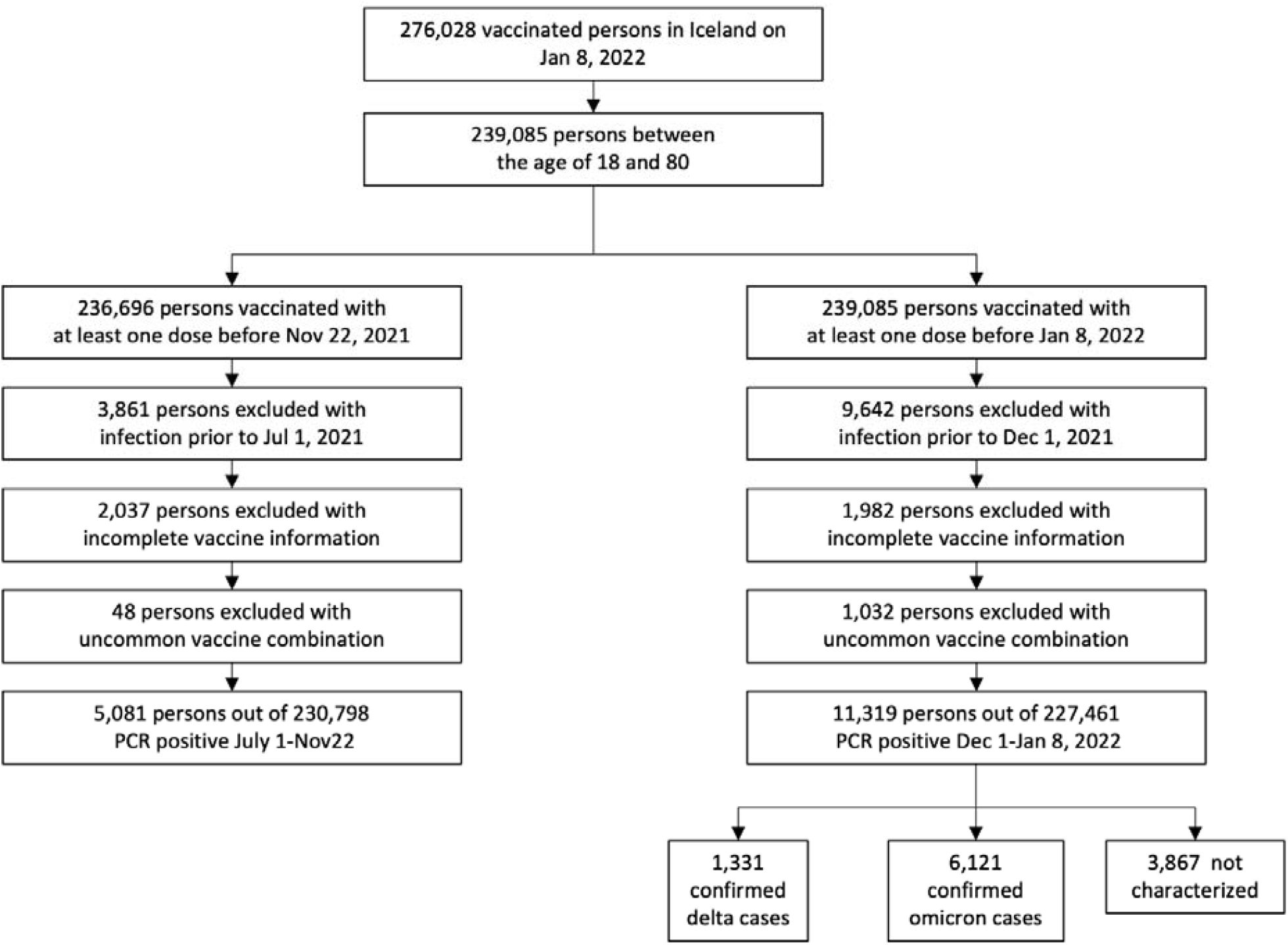
Schematic overview of the selection in the retrospective total population study.

### Antibody and ACE2-Spike binding inhibition levels in *vitro*

To analyze anti-Spike-RBD antibody levels we used recipients of BNT162b2-BNT162b2 as a reference, as it was the most commonly used SARS-CoV-2 vaccine in Iceland (Figure S2).^22^ Ad26.COV2.S recipients had the lowest antibody levels (6.3x lower than BNT162b2-BNT162b2 mRNA recipients, 95% CI: 3.7-10.4x), recipients of ChAdOx1-ChAdOx1 and BNT162b2-BNT162b2 had similar levels (95% CI: 0.6-1.4x) and recipients of mRNA-1273-mRNA-1273 had the highest antibody levels (6.0x higher than BNT162b2-BNT162b2 recipients, 95% CI: 3.8-9.3x) (Figure 1). We did not observe a significant effect of age or sex on antibody levels (95% CI: 0.999-1.03x and 95% CI: 0.49-1.08x, respectively).

The Spike-RBD antibody and ACE2-Spike binding inhibition levels were highly correlated (R=0.82), and the vaccine groups showed a similar separation for both measurements. Both Ad26.COV2.S and ChAdOx1 vaccines elicited lower inhibition levels than BNT162b2 (3.7x, 95% CI: 2.4-5.9x, and 2.8x, 95% CI: 2.0-3.9x, respectively) while mRNA-1273 induced higher inhibition levels (3.3x, 95% CI: 2.5-4.3x).

### Effect of mRNA vaccine booster on antibody and inhibition levels

Of the 106 recruited persons, 99 participants gave their first sample prior to or within four days after receiving the booster (Figure S3 and S4, Table 1), and donated a second sample 50-70 days post-booster, 66 received mRNA-1273 and 33 BNT162b2 as a booster. For comparison, we recruited 59 two-dose BNT162b2 recipients, age and sex matched to the Ad26.COV2.S pre-booster group.

We observed markedly lower antibody levels in the Ad26.COV2.S pre-booster group than the reference group (8.8x, 95% CI: 7.0-11.1x) (Figure 3B, Table 1), whereas, recipients of Ad26.COV2.S-mRNA-1273 showed antibody levels 4.4-fold higher than the BNT162b2-BNT162b2 reference group (95% CI: 3.5-5.7x), and recipients of Ad26.COV2.S-BNT162b2 had 2.6-fold higher levels (95% CI: 1.9-3.5x). It is noteworthy that the Ad26.COV2.S-mRNA-1273 group had higher antibody levels than the Ad26.COV2.S-BNT162b2 group (1.7x, 95% CI: 1.4-2.2x). The mean increase in antibody levels was 36.3-fold for Ad26.COV2.S with booster mRNA-1273 (95% CI: 31-43x) and 26.6-fold for Ad26.COV2.S with booster BNT162b2 (95% CI: 21-34x) (Figure 3C).

**Figure 3.**
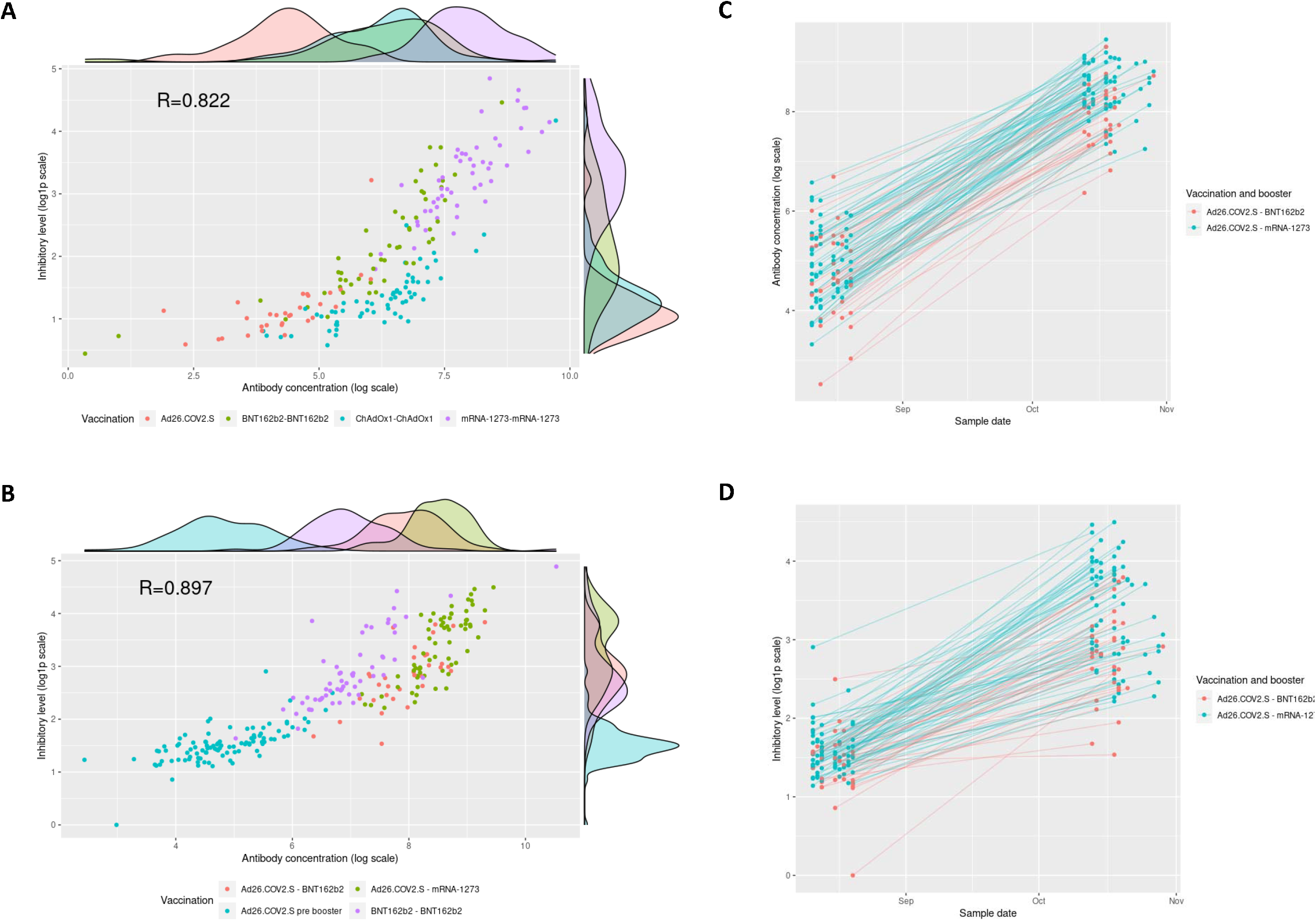
Antibody and inhibition levels by vaccination. A) Anti-Spike-RBD antibody and ACE2-Spike binding inhibition levels across vaccine groups. B) Anti-Spike-RBD antibody and ACE2-Spike binding inhibition levels before and after booster vaccination. C) Anti-Spike-RBD antibody levels in persons vaccinated with Ad26.COV2.S before and after an mRNA booster. D) ACE2-Spike binding inhibition levels in persons vaccinated with Ad26.COV2.S before and after an mRNA booster.

Prior to receiving a booster, Ad26.COV2.S recipients showed 4.7-fold lower ACE2-Spike binding inhibition levels (95% CI: 3.8-6.0x) (Figure 3B, Table 1) than the BNT162b2-BNT162b2 reference group. Ad26.COV2.S-BNT162b2 recipients were indistinguishable from the BNT162b2-BNT162b2 reference group (95% CI: 0.83-1.36x), whereas the Ad26.COV2.S-mRNA-1273 group showed 1.6-fold higher inhibition levels than the BNT162b2 reference group (95% CI: 1.3-2x). The mean increase in inhibition levels was 5.0-fold in mRNA-1273 and 3.0-fold in BNT162b2 booster recipients (95% CI: 4.2-6.0x and 95% CI: 2.3-4.0x, respectively) (Figure 3D). Thus, the increase was 1.6-fold higher in booster recipients of mRNA-1273 than BNT162b2 (95% CI: 1.2-2.4x). We neither observed a significant effect of age or sex on antibody levels (95% CI: 0.98-1.005x and 95% CI: 0.88-1.26x respectively) nor on inhibition levels (95% CI: 0.996-1.02x and 95% CI: 0.80-1.07x respectively). Furthermore, ACE2-Spike binding inhibition levels against nine additional Spike variants, including delta, followed a similar trend with Ad26.COV2.S-mRNA-1273 eliciting the highest levels (Figure S5 and Table S3).

### Age association of anti-Spike-RBD antibody levels after vaccination

We did not detect an effect of age on antibody levels following vaccination, possibly due to a relatively narrow age distribution in each group. Thus, we combined the two 2xBNT162b2 vaccine groups (n=52 and n=59) that were matched for time from vaccination, spanning ages 22 to 68 years (Figure 1A and 1B). We observed that antibody levels decreased with age (−24% per 10 years, 95% CI: 14-32%), (Figure S6) whereas sex and time from vaccination did not show an effect (95% CI: 0.8-1.55x and 95% CI: 0.17-1.47x per month).

### Antibody waning following an mRNA booster dose

Waning of antibody levels and protection following vaccination against SARS-CoV-2 has been reported.^23–26^ For recipients of two doses of ChAdOx1 and one booster dose of mRNA-1273, antibody levels were substantially increased two months after the booster (12x, 95% CI: 8.9-17x) (Figure 4A and 4B). We used measurements obtained 8-154 days after booster to estimate the waning of antibody and inhibition levels and found it to correspond to a half-life of 51 (95% CI: 48-55) and 42 days respectively (95% CI: 38-46)(Figure 4). We noticed that the waning was not consistent with a steady decay, suggesting a plateauing of the antibody levels as has been suggested.^27^ We estimate the half-life of antibody levels to be 42 days (95% CI: 39-45) for the first 100 days, but 93 days (95% CI: 78-116) for the remaining time period.

**Figure 4.**
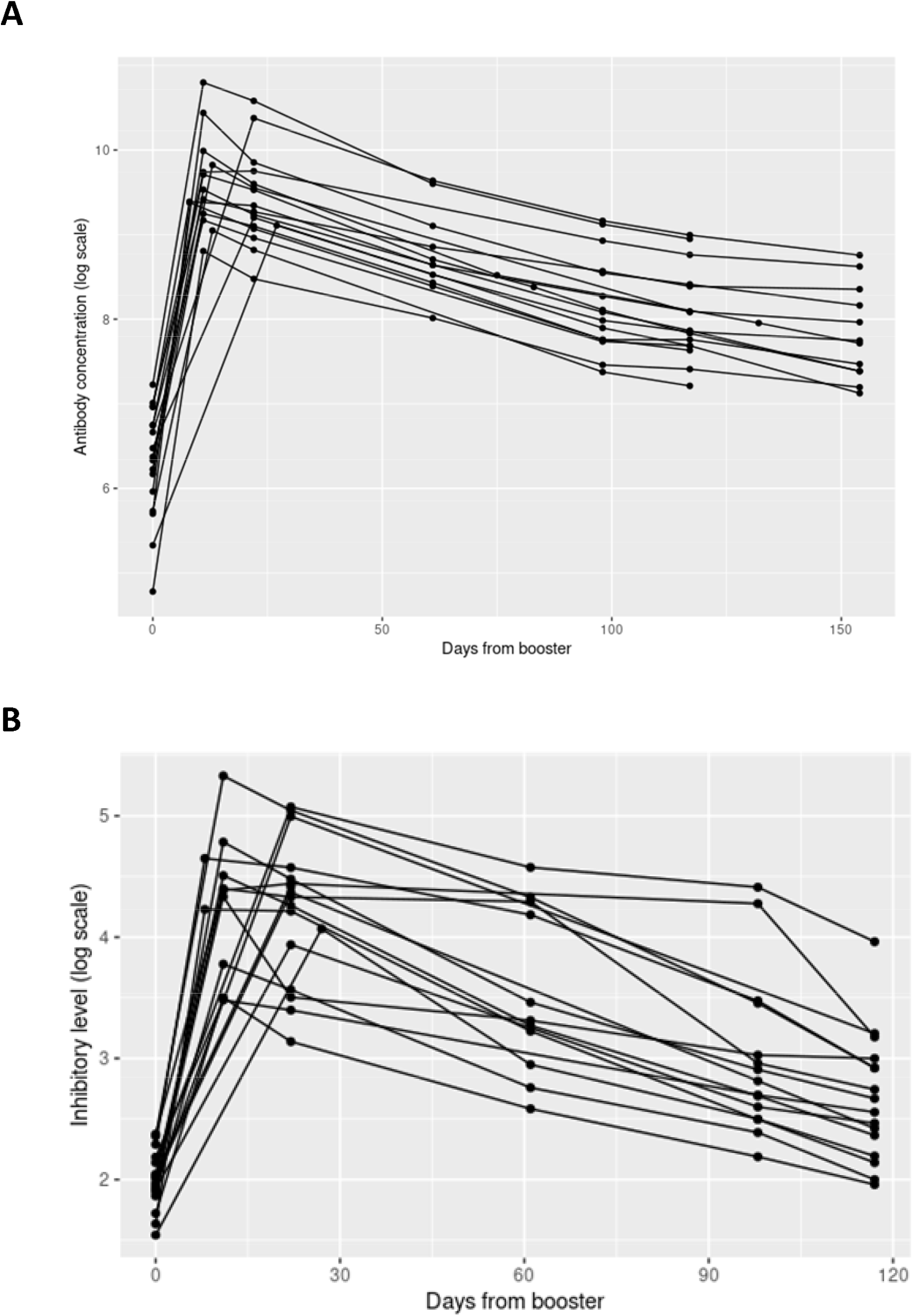
Waning of antibody and inhibitory levels following a booster dose of mRNA-1273. A) Anti-Spike-RBD antibody levels in persons vaccinated with two doses of ChAdOx1 vaccine before and after mRNA-1273 booster vaccination. B) ACE2-Spike binding inhibition levels in persons vaccinated with two doses of ChAdOx1 vaccine before and after mRNA-1273 booster vaccination.

### Effect of vaccination on the risk of COVID-19 diagnosis caused by delta

We restricted the analysis of SARS-CoV-2 infections to the period July 1-November 22, 2021, when a high fraction of the adult population (87%) had received a full vaccination and a peak in infections was dominated by the delta VOC (Figure S1). Figure S7 shows the observed cumulative hazard of SARS-CoV-2 infection for different vaccination schedules. The increase in hazard rates was not constant due to varying incidence of SARS-CoV-2 in the community. Using a Cox proportional hazard model, we regressed out the covariates age, sex and time from vaccination prior to the start date and used BNT162b2-BNT162b2 as a reference group. The adjusted cumulative hazard rate is shown in Figure 5, where the risk is adjusted w.r.t. the effect of covariates. A single Ad26.COV2.S dose was less effective than the BNT162b2-BNT162b2 reference group (Hazard ratio (HR): 2.2, 95% CI: 2.0-2.4), two doses of ChAdOx1 were also less effective (HR: 1.6, 95% CI: 1.43-1.70), whereas two doses of mRNA-1273 were more effective than the BNT162b2-BNT162b2 reference group (HR: 0.8, 95% CI: 0.71-0.91) (Table 2). However, a booster dose of BNT162b2 offered Ad26.COV2.S vaccinees a 1.6-fold higher protection than the BNT162b2-BNT162b2 (HR: 0.62, 95% CI: 0.52-0.73) and 2-fold higher (HR: 0.5, 95% CI: 0.42-0.60) if mRNA-1273 was given as a booster. Similarly, persons who received a third vaccine dose (booster), had a greater protection than the BNT162b2-BNT162b2, or 3.7-fold higher (HR: 0.27, 95% CI: 0.14-0.47) for recipients of BNT162b2-BNT162b2-BNT162b2 and 7.5-fold higher (HR: 0.13, 95% CI: 0.043-0.42) for recipients of ChAdOx1-ChAdOx1 and a BNT162b2 booster.

**Table 2.**
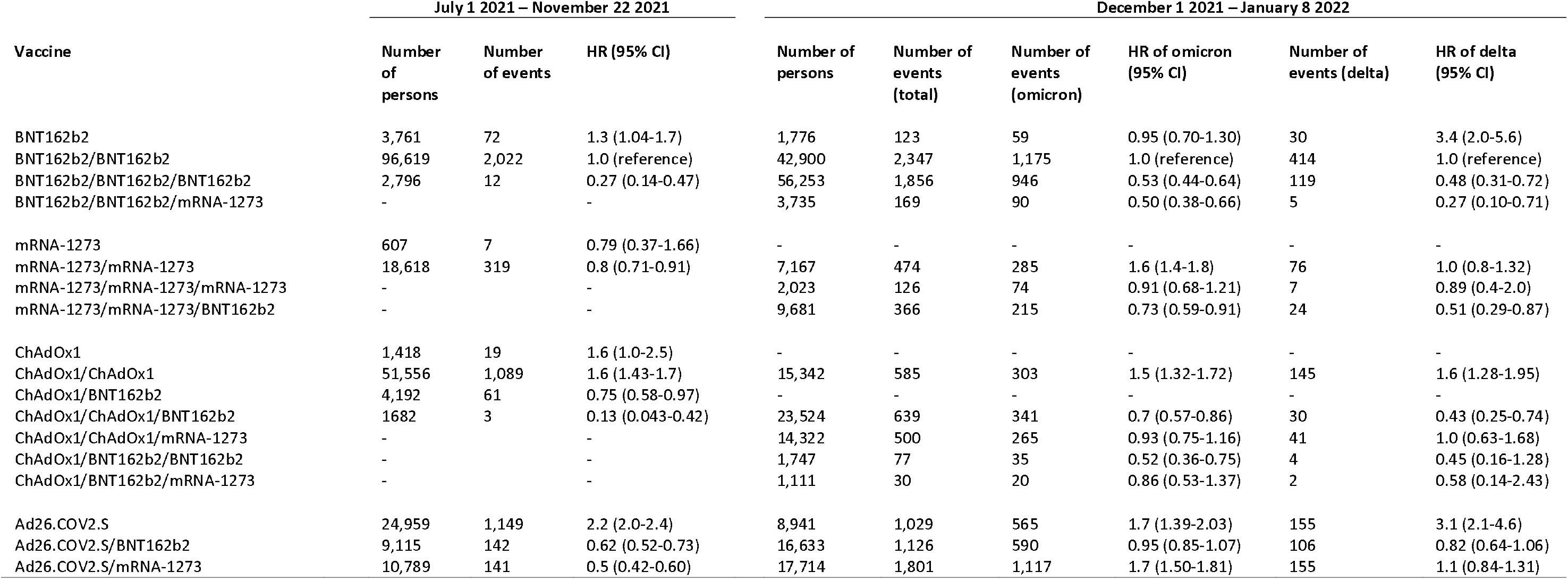
Risk of PCR-confirmed COVID-19 diagnosis by vaccination schedules relative to two dose BNT162b2 vaccination from December 1 2021 – January 8 2022. Number of persons is the average size of each group over the indicated study periods due to vaccinations administered during the study periods. Hazard ratios shown are calculated using BNT162b2/BNT162b2 as the reference group.

**Figure 5.**
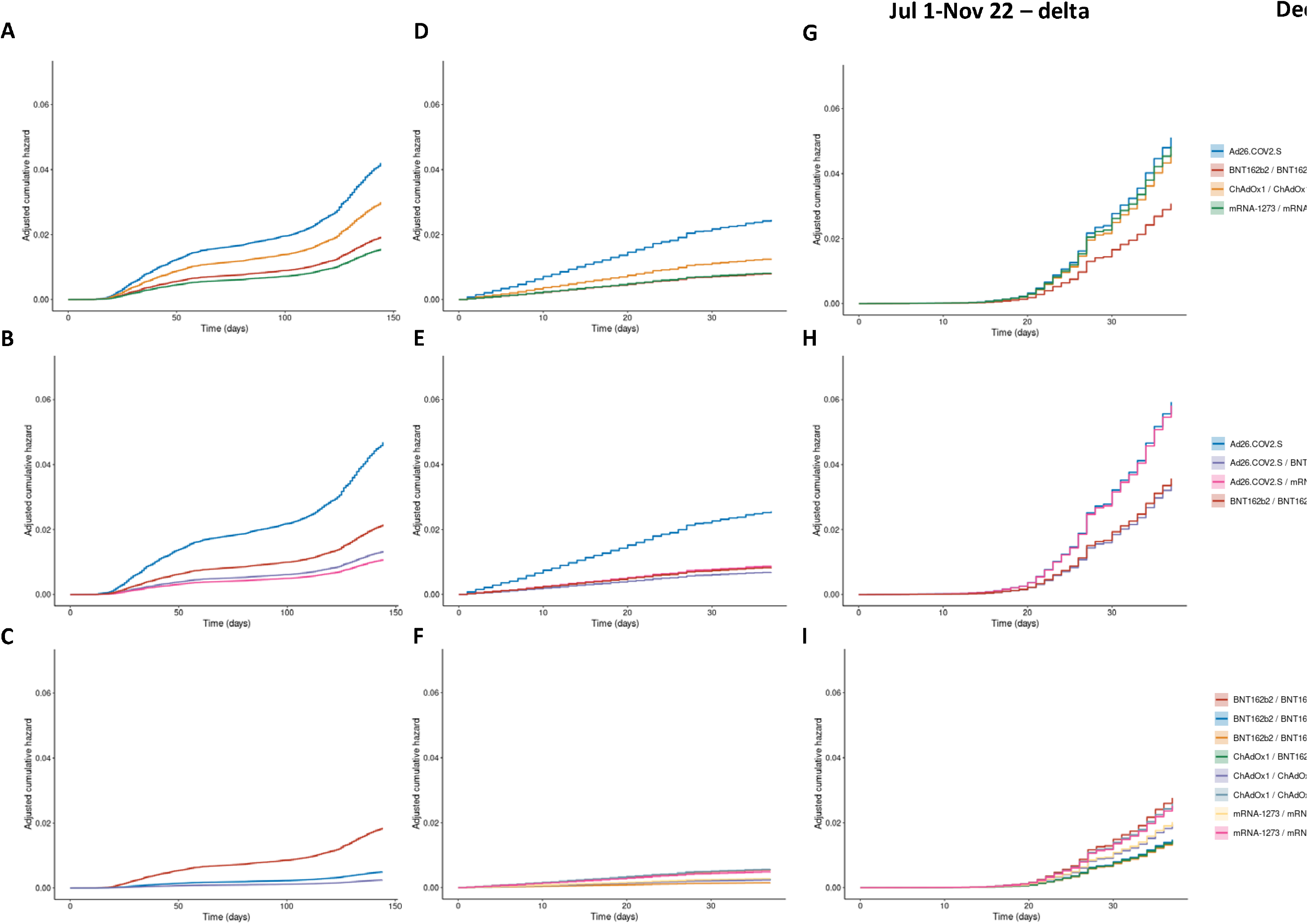
Adjusted cumulative hazard rate of SARS-CoV-2 infections. A-C) Infections caused by the delta variant July 1 – November 22, 2021. D-F) Infections caused by the delta variant December 1, 2022– January 8, 2022. G-I) Infections caused by the omicron variant December 1– January 8, 2022. Top row: Recommended vaccine schedules. Middle row: Comparison of mRNA booster vaccination of Ad26.COV2.S recipients with BNT162b2-BNT162b2 as reference. Bottom row: Comparison of three dose vaccination with BNT162b2-BNT162b2 as reference.

### Effect of vaccination on the risk of COVID-19 diagnosis caused by omicron

To estimate the protection against infection by the newly emerged omicron VOC we restricted our analysis to the period of December 1, 2021 – January 8, 2022, where 90% infections were caused by omicron at the end of the period, with a concomitant increase of more than 1000 PCR-confirmed infections per day (Figure S1). Sequencing of viral samples from infected individuals allowed us to estimate protection against both delta and omicron VOC, and used the 2xBNT162b2 as a reference group (Table 2).

Using the same Cox proportional hazard model, we observed that one dose of BNT162b2 was 3.4-fold less effective against the delta variant than the BNT162b2-BNT162b2 reference group (HR: 3.4, 95% CI: 2.0-5.6) whereas against omicron it was on par with the BNT162b2-BNT162b2 reference group (HR: 0.95, 95% CI: 0.70-1.30). Three doses of BNT162b2 provided a 1.9-fold greater protection against omicron (HR: 0.53, 95% CI: 0.44-0.64) and 2.1-fold greater against delta variant (HR: 0.48, 95% CI: 0.31-0.72) than BNT162b2-BNT162b2. We observed a comparable protection for BNT162b2-BNT162b2-mRNA-1273, as for BNT162b2-BNT162b2-BNT162b2, in comparison with the BNT162b2-BNT162b2 reference group (HR: 0.5, 95% CI: 0.38-0.66, for omicron and HR: 0.27, 95% CI: 0.10-0.71 for delta).

Two doses of mRNA-1273 provided a similar protection against delta as the reference group (HR: 1.0, 95% CI: 0.8-1.32), but 1.6-fold lower protection against omicron (HR: 1.6, 95% CI: 1.4-1.8). An mRNA-1273 booster on top of two doses of the same vaccine, provided similar reduction of infections to that of BNT162b2-BNT162b2 for both variants (95% CI: 0.7-1.2). In contrast, a BNT162b2 booster in the same group provided a 2-fold greater protection against delta (HR: 0.51, 95% CI: 0.29-0.87) than the BNT162b2-BNT162b2 reference group and 1.4-fold greater against omicron (HR: 0.73, 95% CI: 0.59-0.91).

Two doses of ChAdOx1 were inferior against omicron and delta, or 1.5-fold (HR: 1.5, 95% CI: 1.1-1.7) and 1.6-fold (HR: 1.6, 95% CI: 1.3-1.9) respectively, to BNT162b2-BNT162b2. A booster dose of BNT162b2 elicited a reduction of omicron and delta infections, as compared to the reference group, or 1.4-fold (HR: 0.7, 95% CI: 0.57-0.86) and 2.3-fold (HR: 0.43, 95% CI: 0.25-0.74), respectively. In contrast, a booster dose of mRNA-1273 provided a greater protection than two doses of ChAdOx1 alone, but it was not significantly different from the BNT162b2-BNT162b2 reference group (95% CI: 0.6-1.7). One dose of ChAdOx1 followed by two doses of BNT162b2 provided a greater protection against omicron (HR: 0.52, 95% CI: 0.36-0.75) than the reference group, whereas protection against delta was not significantly different (HR: 0.45, 95% CI: 0.16-1.28).

A single dose of Ad26.COV2.S provided 3.1-fold lesser protection against the delta variant (HR: 3.1, 95% CI: 2.1-4.6) and 1.7-fold lesser of omicron (HR: 1.7, 95% CI: 1.4-2.0) than BNT162b2-BNT162b2. A booster dose of BNT162b2, after Ad26.COV2.S, provided a comparable protection as BNT162b2-BNT162b2 against both variants (HR: 0.95, 95% CI: 0.85-1.1, for omicron and HR: 0.82, 95% CI: 0.64-1.1 for delta). Surprisingly, while a booster dose of mRNA-1273 elicited a comparable reduction of delta infections as the BNT162b2-BNT162b2 reference group (HR: 1.1, 95% CI: 0.84-1.31), the protection against omicron was 1.6-fold lower (HR: 1.6, 95% CI: 1.5-1.8).

## Discussion

Based on anti-Spike-RBD antibody levels two months after vaccination we were able to rank the four SARS-CoV-2 vaccines administered by recommended schedules, with one dose of Ad26.COV2.S eliciting the lowest levels and two doses of mRNA-1273 the highest, while two doses of ChAdOx1 and BNT162b2 could not be separated. We observed a good correlation between the levels of antibodies targeting the Spike-RBD and the activity of these antibodies as measured by inhibition of ACE2-Spike binding. Despite this strong correlation, antibody levels alone could not distinguish between two dose ChAdOx1 and BNT162b2 vaccine recipients, whereas they were clearly separated by inhibition levels, with BNT162b2 inducing greater inhibition. The difference between two doses of ChAdOx1 and BNT162b2 was also reflected in their difference in reducing SARS-CoV-2 infections in the Icelandic population. This demonstrates a benefit of measuring inhibitory activity of antibodies, in addition to their levels. Furthermore, this ordering based on inhibitory activity is consistent with their protective capacity against SARS-CoV-2 infections, with the mRNA vaccines providing the greatest protection.

In light of waning immunity over time^23–26^, health authorities in many countries are now recommending booster vaccinations.^12^ The relatively low antibody levels and protective capacity in recipients of one dose of Ad26.COV2.S motivated us to investigate their levels following an mRNA booster, due to the relatively high proportion of Icelanders that received Ad26.COV2.S. All Ad26.COV2.S recipients who received a booster dose of either BNT162b2 or mRNA-1273 showed not only a robust elevation in antibody and inhibition levels, but outperformed the two dose BNT162b2 reference group with mRNA-1273 booster eliciting the greater effect. We were therefore interested to see if this translated into greater protection against SARS-CoV-2 infection. Indeed, both combinations, Ad26.COV2.S-BNT162b2 and Ad26.COV2.S-mRNA-1273, showed greater protection than the two dose BNT162b2 reference against SARS-CoV-2 infection. Given the relatively low protection by a single dose of Ad26.COV2.S, the protection elicited by Ad26.COV2.S-BNT162b2 and Ad26.COV2.S-mRNA-1273 is striking. This seems to be consistent with emerging data suggesting that a vaccination schedules that mix two vaccine types, adenoviral and mRNA, elicit a greater immune response than that with only one type^28^, including better T-cell immunity.^29,30^ These data are in strong support of the booster strategy implemented in Iceland, where recipients of a single dose of Ad26.COV2.S were moved from having the highest risk of infection out of those following the recommended vaccine schedule, to having among the lowest risk.

The complete nationwide data on vaccinations and qPCR diagnosis allowed us to assess the reduction of SARS-CoV-2 infection by other booster vaccinations not included in our smaller recruited groups. We observed that an additional third dose of BNT162b2 greatly decreased the risk of being infected in agreement with published data.^7,8,15^ Similarly, recipients of two doses of ChAdOx1 who received an additional dose of BNT162b2 diverged from those who received two doses of BNT162b2, with more reduction of infections comparable to three doses of BNT162b2. This constitutes a striking turn of events as two doses of ChAdOx1 elicited lower protection than two doses of BNT162b2. These data mirror the great increase in antibody and inhibition levels observed when recipients of two doses of ChAdOx1 received a booster of mRNA-1273.

The extremely rapid spread of the omicron VOC and emerging data on vaccine effectiveness both support a lower protection than against the delta variant^16^. Our data confirm that a booster approach is indeed effective in terms of reducing the risk of being infected by omicron. While mRNA-1273 was effective against infection by delta, our data support a much poorer performance against omicron. This was not only apparent for recipients of two doses of mRNA-1273, when they were boosted with either BNT16b2 or mRNA-1273, but also for those originally vaccinated with two doses of ChAdOx1 or one dose of Ad26.COV2.S. Despite this, a booster dose of mRNA-1273 provided a similarly increased protection to that of BNT162b2 against both the delta and omicron VOCs, in recipients of two doses of BNT162b2. Importantly, both ChAdOx1 and Ad26.COV2.S retained their relative ordering, providing far lesser protection against both variants compared with two doses of BNT162b2. It is therefore encouraging that a single dose of an mRNA vaccine elevates the protection against both VOCs at least up to the level of two doses of BNT162b2. It is clear though that additional booster doses will be needed for recipients of Ad26.COV2.S.

It must be emphasized that although our data demonstrate a greater protection against omicron, following a booster vaccination compared to two doses of BNT162b2 that serves as a reference, we do not have data on the relative protection compared to those unvaccinated. This comparison is however, less relevant for a country like Iceland where 91% of inhabitants 12 years or older have now been fully vaccinated. The observational nature of the population study presents limitations to consider. There is a confounding factor between age and time of vaccination, namely the first to receive vaccination were front-line workers and the elderly. Further, the vaccine received correlates with age and gender. For the analyses of booster vaccinations the group sizes differ, which is accounted for in the statistical analysis but selection bias could generate statistical artifacts. The key strength of this study is the complete nationwide coverage, in terms of the high fraction of the population vaccinated, the sheer number of PCR tests conducted and sequencing of viral isolates.

The mixed vaccination approach in Iceland provided a unique opportunity to investigate the effects of various vaccination schedules and booster combinations within a single population. Our data clearly demonstrate and confirm the major benefit of mRNA booster vaccinations against SARS-CoV-2 infections, both against the delta and omicron VOCs, regardless of the vaccine type given in the primary series. We observe a great increase in antibody level and inhibitory activity, consistent with our estimates of the effectiveness of protection against SARS-CoV-2 infections by mRNA booster vaccinations at a population level.

## Methods

### Exposures and outcomes in the total population cohort study

We used vaccination status as exposure variable, where each vaccine combination creates a separate group at each point in time. Persons who received a vaccine dose during the study period changed their exposure in a time dependent manner. During the first time period we defined the outcome as a positive PCR test, regardless of symptoms and severity. For the second time period, we used PCR and sequencing data to classify positive tests into delta or omicron outcomes. Persons who received a positive test, that could not be classified, did not receive an outcome.

### Antibody measurement

Pan-Ig antibody levels against SARS-CoV-2 Spike-RBD in serum samples was quantified using Elecsys anti-SARS-CoV-2 S (Roche Diagnostics #09289267190) on Cobas e601. The linear range of the assay is 0.4-250 U/mL. Samples were measured undiluted and if the concentration was higher than 250 U/mL, they were diluted 1:10, until concentration was in the linear range.

### Inhibition measurement

Inhibition of ACE2 binding to the Spike protein by serum samples was measured using a multiplexed immunoassay (Meso Scale Diagnostics, LLC #K15466U, #K15436U) as a surrogate for neutralizing capacity of antibodies in serum, which are known to correlate.^19^ The manufacturer’s protocol was followed. Samples were diluted 1:50 and 1:500. Following a 1 hour incubation, Sulfo-tag labelled ACE2 was added and incubated for 1 hour. Plates were read using a MESO® SECTOR S600 Reader.

SARS-CoV-2 Spike monoclonal neutralizing antibody was used to generate a calibration curve. To calculate neutralizing antibody concentrations (in units/mL), signals were backfitted to the calibration curve.

### Variant classification

The viral genome of all positive samples were sequenced at deCODE genetics using Illumina or ONT technologies (Supplementary methods). For the most recent infections, where sequencing was not yet available, we classified positive samples into delta or omicron based on S-gene target failure on the Taqman assay, if available.

### Statistical analysis

A linear model was used to calculate the effect of covariates on antibody concentration or inhibition levels. For antibody concentrations, all values were transformed using the natural logarithm (ln) and for inhibition levels values were transformed using the ln of one plus.

We used a Cox proportional hazards regression model to estimate the vaccine effectiveness w.r.t. a reference group as estimated using the hazard ratio. The effect of vaccine combinations are estimated with a time varying covariate to account for vaccinations that occur within the study window. Time from vaccination was taken as a covariate for those persons vaccinated before the study window. We regressed sex and age as covariates and used a penalized splines curve to regress out the effect of age. For any vaccine dose that occurred within the study window, a 14-day lag time was introduced to account for the time until a full effect is observed.

The R statistical software, version 3.6.3 was used for all statistical analysis.^20^ The survival package in R was used for all survival analysis.^21^ Confidence intervals reported were not corrected for multiple testing.

## Data Availability

All sequences used in this analysis will be made available in the European Nucleotide Archive (ENA) under accession number PRJEB44803 (https://www.ebi.ac.uk/ena/browser/view/PRJEB44803)

## Ethical consideration

The study was approved by the National Bioethics Committee of Iceland (approval no. VSN-21-072), after review by the Icelandic Data Protection Authority (DPA). Participants who donated blood signed informed consent. The personal identities of participants were encrypted using a third-party system approved and monitored by the DPA.

## Author contributions

G.L.N., P.M., I.J and K.S. designed the study. G.L.N. and P.M. conducted the analysis and G.H.H., A.G. and D.F.G. assisted with the analysis. I.J. performed recruitment. O.T.M. performed sequencing. K.G: performed laboratory work. G.L.N., P.M., I.J, K.S., D.F.G, K.G., T.A.O., M.K., P.S., U.T. interpreted the results. All authors contributed to the final version of the manuscript.

