## Supplementary Appendix for "Effect of booster vaccination against Delta and Omicron variants in Iceland"

### Supplementary methods

#### Sequencing

The viral genomes of all positive PCR samples were sequenced at deCODE genetics, the methods have been previously described^1^ and are reproduced here for the benefit of the reader. Reverse transcription and multiplex amplicon PCR were performed on the basis of information provided by the Artic Network initiative (https://artic.network/) to generate complementary DNA and sequencing libraries. Samples were sequenced using either Illumina or ONT sequencing technologies. Illumina sequencing was performed using MiSeq sequencers (MiSeq v2 reagent kits) with 2 x 150 cycle paired-end reads, with up to 48 multiplexed samples per run. Samples for ONT were multiplexed using native barcodes and sequenced using either GridION or PromethION flowcells, version R.9.4.1. Details are provided in the Supplementary methods.

#### RNA extraction

Viral RNA samples were extracted from swabs (96 samples per 40 min run) using the Chemagic Viral RNA kit on the Chemagic360 instrument from Perkin Elmer, with 300/100 µL input/output volume(s), respectively. Each step in the workflow was monitored using an in-house LIMS (VirLab) with 2D barcoding (Greiner, 300 µL tubes) of all extracted positive samples.

#### Reverse transcription and PCR amplification

Reverse transcription (RT) and multiplex PCR was performed based on information provided by the Artic Network initiative (https://artic.network/) to generate cDNA. In short, extracted viral RNA was pre-incubated at 65 °C for 5 min in the presence of random hexamers (2.5 µM) and dNTP´s (500 µM). Sample cooling on ice was then followed by RT using SuperScript IV (ThermoFisher) in the presence of DTT (5 mM) and RNaseOUT inhibitor (Thermo Fisher) for 10 min at 42°C, followed by 10 min at 70 °C. Multiplex PCR of the resulting SARS-CoV-2 cDNA was performed using a tiling scheme of primers, designed to generate overlapping amplicons of approximately 400 bp (Supplementary table 6). Two PCR reactions were performed for each sample using primer pools A and B, respectively (Table S1). PCR amplification was done using the Q5® Hot Start High-Fidelity polymerase (New England Biolabs) with primers at 1 µM concentration. The reactions were performed in an MJR thermal cycler with a heated lid at 105 °C, using 35 cycles of denaturation (15 sec at 98 °C) and annealing/extension (5 min at 65 °C). The resulting PCR amplicon pools A and B were combined, purified using 1X Ampure XP magnetic beads (Beckman Coulter) and quantified using the Qubit dsDNA high-sensitivity assay kit (Thermo Fisher).

#### Sample preparation for ONT Sequencing

Pooled amplicons (50-400 ng) were prepared for ONT sequencing using native barcoding with up to 24 barcodes. The first step in the procedure was end-repair using NEBNext Ultra II End Prep Reaction Buffer and Enzyme mix, respectively (NEB/E7546L). Samples (50 µL) were incubated in an MJR thermocycler at 20 °C for 10 min, followed by 10 min at 65 °C and cooling at 4 °C. Following AMPure XP clean-up (1X), the native barcodes (NBD104/NBD114) were ligated by mixing 20 µL purified sample, 2.5 µL barcode and 25 uL blunt/T4 ligase mastermix (NEB/M0367L). The mixture was incubated at room temperature on a rotator mixer for 10 min. Samples were purified by AMPure XP and quantified using the Qubit dsDNA high-sensitivity assay kit. Samples were pooled appropriately based on yield and/or Ct values from previous RT-qPCR measurements, with up to 24 samples/pool. Pooled DNA samples (up to 90 uL) were next incubated on a rotator mixer with 5 uL adapter mix II (AMII/ONT), 20 uL NEBNext Quick ligation reaction buffer and 10 uL Quick T4 DNA ligase (NEB/E6056L) for 10 min at room temperature followed by AMPure clean-up and bead washing with 2x120 µL of short fragment buffer (SFB/ONT). Final pooled sample libraries were resuspended in 35 uL of EB buffer and used immediately for sequencing.

#### ONT Sequencing

Pooled libraries were sequenced on either GridION X5 or PromethION sequencers, using the MinION (R9.4.1/FLO-MIN106D) and PromethION (R9.4.1/FLO-PRO002) flowcells, respectively. Following priming of the flowcells, the libraries were mixed with sequencing buffer (SQB) and loading beads (LB) and loaded onto the flowcells in the appropriate volume depending on flowcell type (75 µL or 150 µL). Sequencing runs were performed using on-board, high-accuracy basecalling (Guppy) with generation of FASTQ files for each barcode. Data acquisition varied from 1-4 hrs depending on the number of samples/pool and run performance. At least 100K reads were acquired per sample (barcode).

#### Analysis of sequence data

We aligned amplicon sequences to the reference genome of the SARS-CoV-2 (GenBank number, NC_045512.2). For Illumina sequences we used the latest Burrows–Wheeler Aligner (BWA-MEM) and variants were called with sequencing utilities bcftools as previously described.^2^ For ONT sequences, sequence alignment and variant calling was performed using the Artic Network pipeline (https://artic.network/) with default parameters.

#### SARS-CoV-2 testing

To assess the variant of PCR positive samples where sequencing was not yet available, we used SGTF (Spike Gene Target Failure) property of the omicron variant (B.1.1.529) on the TaqPath™ COVID-19 CE-IVD RT-PCR Kit (Catalog number: A48067, ThermoFisher), otherwise the analysis remained unchanged^2^.

### Supplementary figures

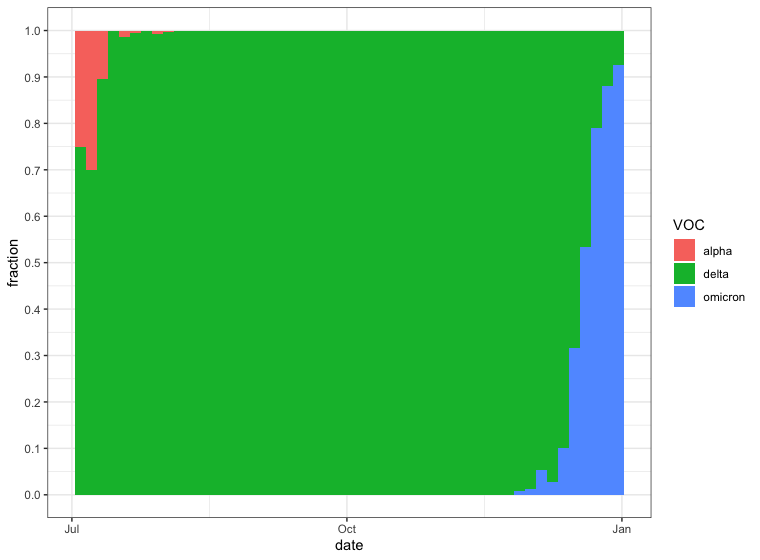

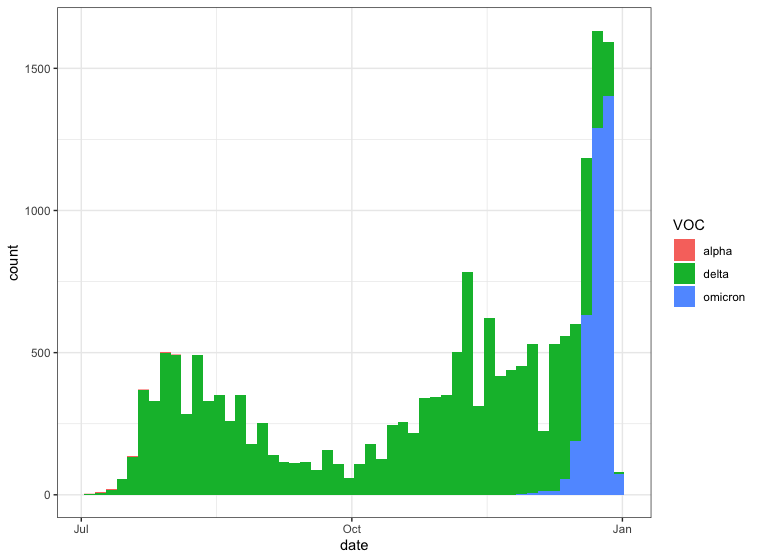

**A**

**B**

Figure S1 A) Fraction of SARS-CoV-2 infections by variant of concern as determined using sequencing. B) Absolute number of SARS-CoV-2 infections by variant. Shown is the time period July 1, 2021-January 8, 2022.

Figure S2 Age and sex distribution of primary schedule vaccination of the four vaccines deployed in Iceland by November 30th 2021.

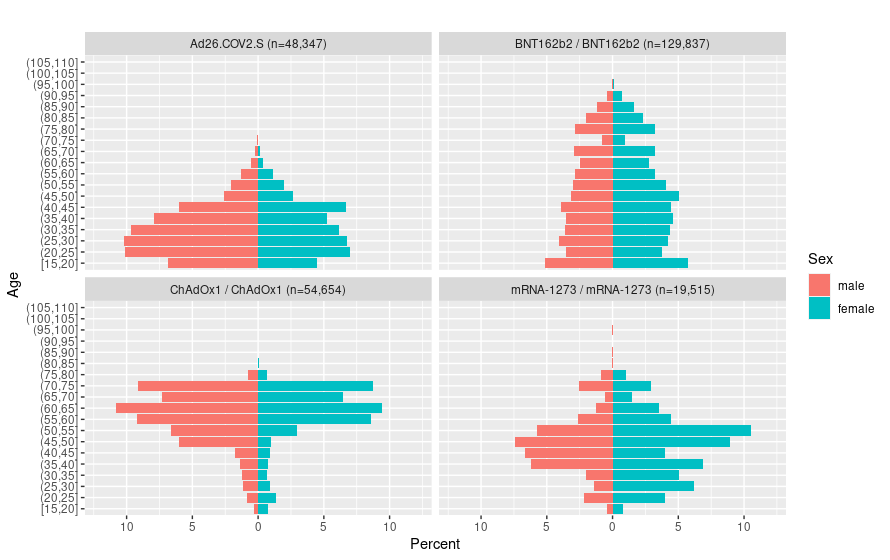

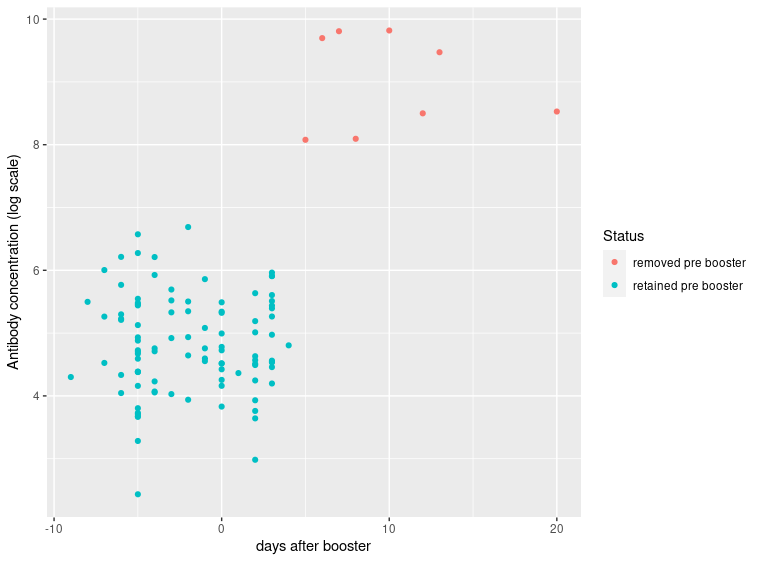

Figure S3 Anti-Spike-RBD antibody levels by number of days before or after booster dose. Red dots show samples removed from further analysis. Eight persons (red) gave their first sample 5 days or more after the mRNA booster and their antibody levels were considerably higher than in the pre-booster group (65x, p<2x10^-16^). However, two months after the booster their antibody levels had lowered and were indistinguishable from the other samples in the post booster group (p=0.44).

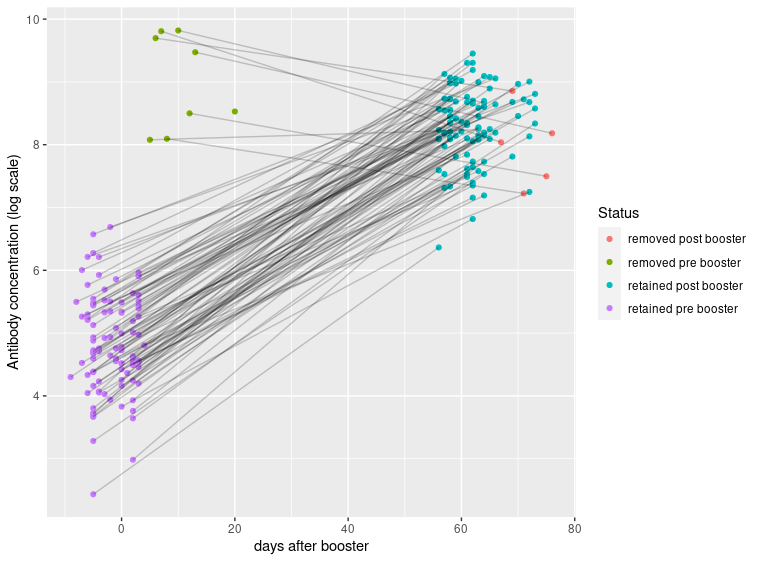

Figure S4 Anti-Spike-RBD antibody levels by number of days before or after booster dose in all persons with two samples.

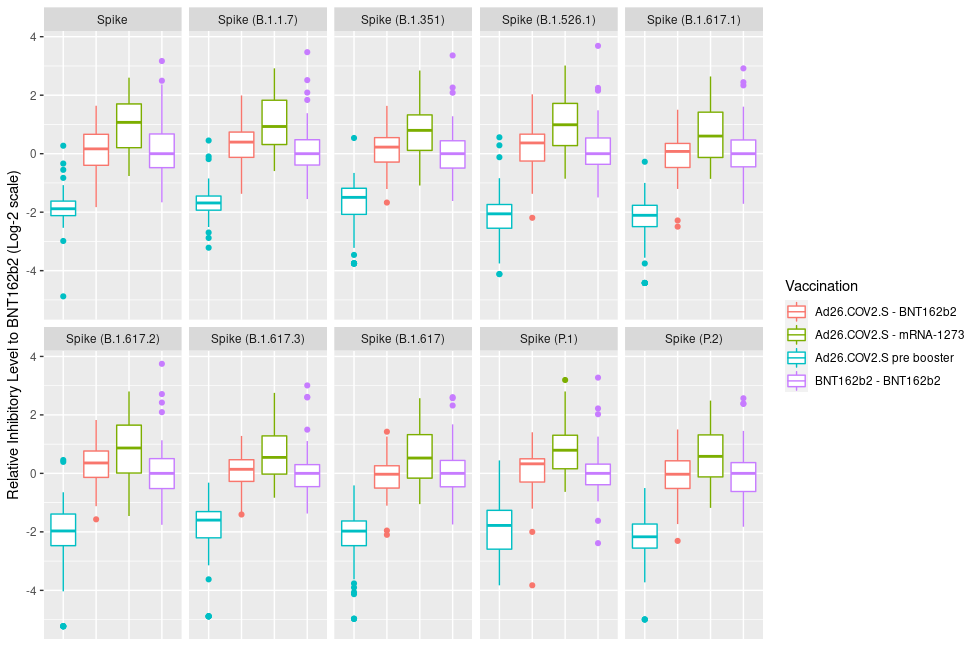

Figure S5 ACE2-Spike binding inhibition levels across Spike protein variants in persons vaccinated with Ad26.COV2.S before and after receiving mRNA booster vaccination. Data normalized to the two dose BNT162b2 reference group. We measured levels of ACE2-Spike binding inhibition in serum using nine additional Spike variants, including delta. The ACE2-Spike binding inhibition levels prior to booster were lower in recipients of Ad26.COV2.S than two dose BNT162b2 for all Spike variants tested, as observed for the original Spike (Table S1). Similarly, the Ad26.COV2.S-mRNA-1273 group showed the highest relative ACE2-Spike binding inhibition levels post-booster, whereas Ad26.COV2.S-BNT162b2 did not show a statistically significant difference from the two-dose BNT162b2 mRNA reference group, irrespective of Spike variants.

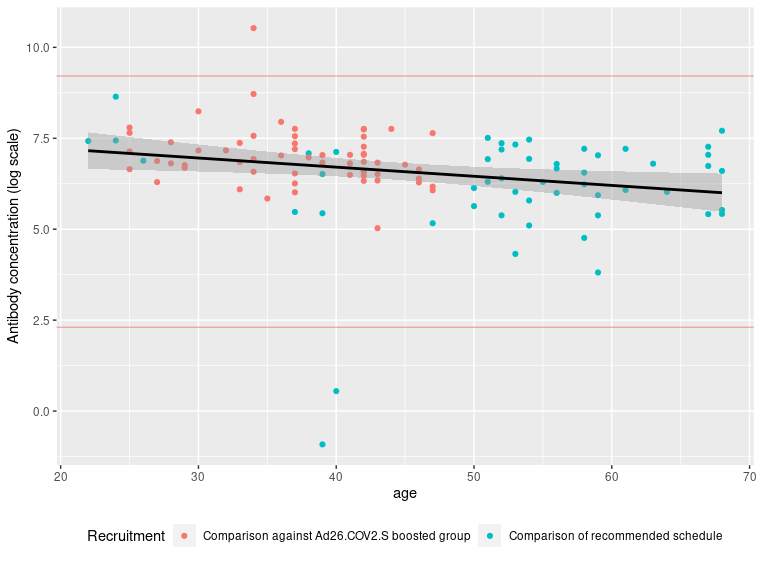

Figure S6 Anti-Spike-RBD antibody levels two dose BNT162b2 vaccinees: the reference group for Ad26.COV2.S receiving mRNA booster (red, N=52)(Figure 2B), reference group for the recommended two dose schedules (cyan, N=59)(Figure 2A). Red lines show the limits of outliers removed at 10 and 10000 U/ml.

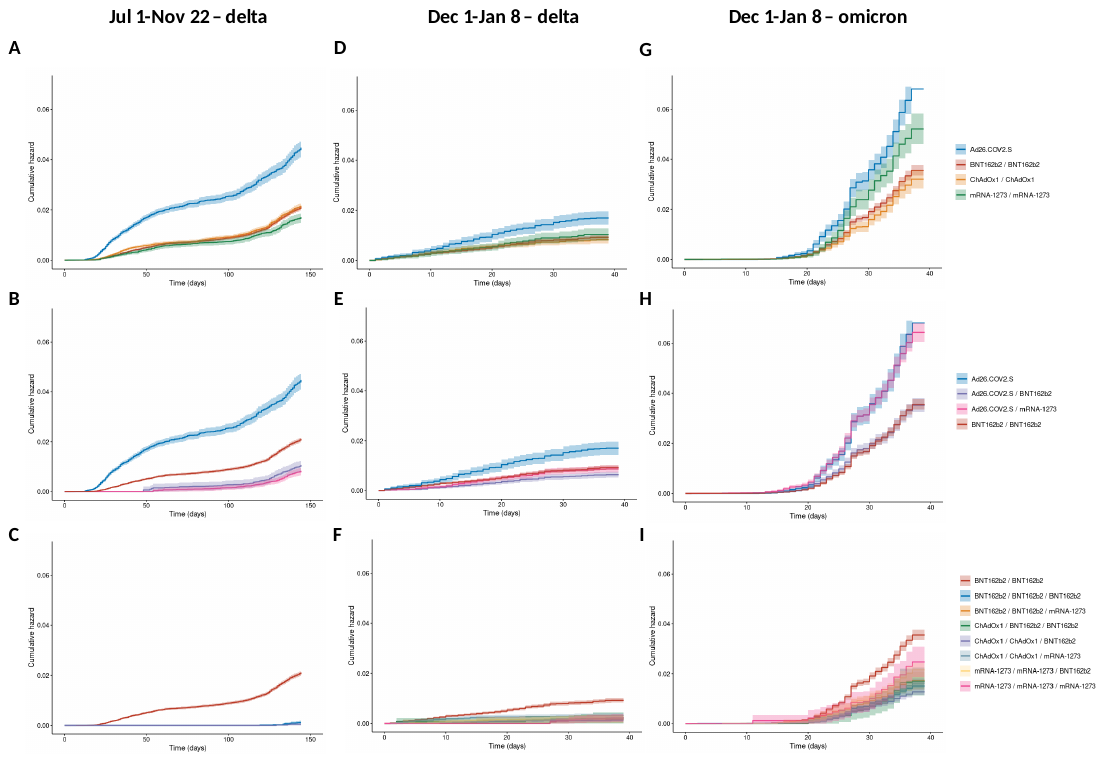

Figure S7. Kaplan-Meier plots comparing SARS-CoV-2 infections. A-C) Infections caused by the delta variant July 1 – November 22, 2021. D-F) Infections caused by the delta variant December 1, 2022– January 8, 2022. G-I) Infections caused by the omicron variant December 1, 2022– January 8, 2022. Top row: Recommended vaccine schedules. Middle row: Comparison of mRNA booster vaccination of Ad26.COV2.S recipients with 2xBNT162b2 as reference. Bottom row: Comparison of three dose vaccinations with 2xBNT162b2 as reference.

### Supplementary tables

Table S1 Demographics for the ChAdOx1- ChAdOx1-mrna-1273 recipients.

|  | Overall |
| --- | --- |
| Number | 17 |
| Female (%) | 3 (18%) |
| Age | 58.4 years |
| Time from the ChAdOx1-ChAdOx1 vaccination | 101 days |

**Table S2** Demographics of vaccination groups on January 8, 2022.

|  | January 8 2022 | | | | |
| --- | --- | --- | --- | --- | --- |
| **Vaccine** | **Number of  persons** | **Age (IQR)** | **Female (%)** | **Days between dose number 2 and 3** | Days between dose number 1 and 2 |
| BNT162b2 | 1,952 | 39 (27-48) | 49.3 |  |  |
| BNT162b2/BNT162b2 | 28,468 | 36.5 (25-43) | 52.7 |  | 25.1 |
| BNT162b2/BNT162b2/BNT162b2 | 69,206 | 49.3 (36-64) | 54.2 | 185.7 | 22.0 |
| BNT162b2/BNT162b2/mRNA-1273 | 5,892 | 43.8 (34-52) | 62.5 | 169.4 | 22.5 |
| mRNA-1273/mRNA-1273 | 5,131 | 39.5 (30-47) | 52.8 |  | 30.8 |
| mRNA-1273/mRNA-1273/mRNA-1273 | 3,403 | 50 (43-54) | 52.7 | 171.3 | 27.7 |
| mRNA-1273/mRNA-1273/BNT162b2 | 10,482 | 49 (38-57) | 65.8 | 190.8 | 28.3 |
| ChAdOx1/ChAdOx1 | 6,896 | 52.4 (44-63) | 41.0 |  | 69.1 |
| ChAdOx1/ChAdOx1/BNT162b2 | 29,219 | 61.4 (56-72) | 48.2 | 152.9 | 67.8 |
| ChAdOx1/ChAdOx1/mRNA-1273 | 17,148 | 59.5 (56-64) | 36.0 | 155.5 | 60.6 |
| ChAdOx1/BNT162b2/BNT162b2 | 2,263 | 45.5 (32-58) | 70.5 | 169.3 | 74.8 |
| ChAdOx1/BNT162b2/mRNA-1273 | 1,218 | 56.9 (52-62) | 24.9 | 153.7 | 60.7 |
| Ad26.COV2.S | 8,377 | 31.8 (24-37) | 38.2 |  |  |
| Ad26.COV2.S/BNT162b2 | 17,670 | 34.8 (26-43) | 39.6 |  | 86.8 |
| Ad26.COV2.S/mRNA-1273 | 18,233 | 32.5 (24-40) | 46.9 |  | 73.3 |

Table S3 ACE2-Spike binding inhibition levels relative to the original Spike protein across Spike variants for the different vaccination groups. Vaccine groups within each variant assay were normalized by the median value of the BNT162b2-BNT162b2 reference group. The estimate of the relative effect and confidence intervals are calculated using the normalized value against a null distribution.

| Spike variant | BNT162b2 - BNT162b2 reference group (95% CI) | Ad26.COV2.S  pre-booster (95% CI) | Ad26.COV2.S – BNT162b2 (95% CI) | Ad26.COV2.S – mRNA-1273 (95% CI) |
| --- | --- | --- | --- | --- |
| Spike | 1.2 (1.0-1.4) | 0.23 (0.19-0.28) | 0.94 (0.74-1.2) | 1.6 (1.3-2.0) |
| Spike (B.1.1.7) | 1.1 (0.98-1.3) | 0.28 (0.24-0.33) | 1.1 (0.86-1.4) | 1.8 (1.5-2.2) |
| Spike (B.1.351) | 1.1 (0.92-1.2) | 0.29 (0.24-0.35) | 1.05 (0.83-1.3) | 1.6 (1.3-2.0) |
| Spike (B.1.526.1) | 1.1 (0.98-1.3) | 0.21 (0.17-0.25) | 1.04 (0.8-1.3) | 1.8 (1.4-2.2) |
| Spike (B.1.617.1) | 1.1 (0.93-1.3) | 0.19 (0.16-0.23) | 0.89 (0.69-1.1) | 1.5 (1.2-1.8) |
| Spike (B.1.617.2) | 1.1 (0.91-1.3) | 0.22 (0.17-0.27) | 1.1 (0.83-1.5) | 1.7 (1.4-2.2) |
| Spike (B.1.617.3) | 1.1 (0.91-1.2) | 0.26 (0.21-0.31) | 1.0 (0.78-1.3) | 1.5 (1.2-1.8) |
| Spike (B.1.617) | 1.1 (0.92-1.2) | 0.21 (0.17-0.26) | 0.87 (0.67-1.1) | 1.4 (1.2-1.8) |
| Spike (P.1) | 1.1 (0.9-1.2) | 0.23 (0.19-0.29) | 0.96 (0.73-1.3) | 1.6 (1.3-2.0) |
| Spike (P.2) | 1.0 (0.88-1.2) | 0.21 (0.17-0.25) | 0.93 (0.73-1.2) | 1.5 (1.2-1.8) |
